# The Prognostic Value of PD-1/PD-L1 Expression on Tumor Cells and Tumor-Infiltrating Immune Cells in Patients with Colorectal Cancer: a Systematic Review and Meta-Analysis Protocol

**DOI:** 10.1101/2021.03.09.21253153

**Authors:** Fateme Abedini, Saeedeh Salehi, Leila Janani, Monireh Mohsenzadegan

## Abstract

**Aim:** Colorectal cancer (CRC) is one of the most common cancers in the world. However, the role of immune checkpoint molecules, especially Programmed cell death protein 1 (PD-1) and Programmed cell death-ligand 1 (PD-L1), in the progression of CRC remains unclear. This systematic review and meta-analysis will investigate the prognostic significance of PD-1/PD-L1 expression on tumor-infiltrating immune cells and tumor cells in patients with colorectal cancer.

**Methods:** This protocol has been prospectively registered in the PROSPERO (registration NO. CRD42020156233). A comprehensive electronic search on PubMed/MEDLINE, Scopus, Web of Science (WOS), Embase and ProQuest will be conducted using a combination of MeSH terms of “programmed cell death 1”, “programmed cell death ligand 1”, “colorectal” and “cancer” between 1 January 1990 and 31 March 2021 with no language limitation. Two independent reviewers will perform study selection, data extraction, and risk of bias assessment. The Newcastle-Ottawa Scale (NOS) for cohort studies will be used to assess the risk of bias. In the case of sufficient data, either random or fixed-effect models, will be used for meta-analysis. Statistical heterogeneity will be evaluated by 𝒳^2^ test with the I^2^ statistic. “Funnel plot”, “Begg’s statistical test”, and “Egger’s statistical test and graph” will be used to assess publication bias. Stata V.13 software will be employed for meta-analysis.

**Results and conclusion:** According to the meta-analysis of the aggregated data from the relevant primary studies, the relationship between expression of PD-1/PD-L1 and prognostic parameters, including progression-free survival and overall survival, will be reported. The results of the current study will be published in a peer-reviewed journal.

## 1. Introduction

Colorectal cancer (CRC) is the second most common cancer in men and women after prostate and breast cancer, respectively. It is also the second lethal cancer after lung cancer in the world (1). The 5-year survival rate in patients with CRC is about 64%. However, it relates to several factors, such as the stage of cancer, age, and overall health. For instance, 14% of people who have distant metastases have 5-year survival (2). Although the rate of death from CRC has been decreasing due to the improvement of screening and treatment, it remains a leading cause of death worldwide for its poor prognosis, especially in the late stages. Therefore, it is crucial to identify biomarkers to improve patient’s outcome.

The progression of CRC is influenced by complex interactions between tumor microenvironment (TME) and tumor cells. TME employs many mechanisms to suppress the immune system (3); one of them is programmed cell death protein 1/programmed cell death-ligand 1 (PD-1/PD-L1) axis, which affects the balance between immune resistance and tumor immune surveillance (4).

PD-1 (CD279) is an inhibitory co-receptor expressed on the surface of activated immune cells, including dendritic cells, macrophages, natural killer (NK) cells, and tumor-infiltrating lymphocytes (TILs; e.g., T cells, and B cells) (5). PD-L1 (CD274) - the primary ligand of PD-1 - is expressed by the immune system or cancer cells (6). Immune cells, such as activated lymphocytes (T, B, and NK cells) express PD-L1 on their surface (3). The high expression of PD-L1 on TILs or tumor cells leads to T cell exhaustion and promotes tumor progression (4).

The interaction between PD-1 and PD-L1 sends inhibitory signals that block the production of cytotoxic mediators required for the killing of tumor cells and also reduces the proliferation of T cells (7, 8).

The relation between unusual expression of PD-L1 and prognosis or response to the treatment has been observed in several cancers. For example, the relationship between PD-L1 expression and prognosis has been detected in different solid tumors, including urothelial carcinoma, non-small cell lung cancer (NSCLC), and ovarian cancer (9-11). Furthermore, several studies have investigated the relationship between the expression of PD-1/PD-L1 and survival in patients with CRC; some of them have shown that patients with overexpression of these molecules have better survival than others (12, 13), but other studies have illustrated opposite results or no relationship (14, 15).

Due to these controversies, several systematic reviews and meta-analyses have been conducted in order to evaluate the prognostic significance of PD-L1 expression in patients with CRC and other cancers. Two meta-analyses have been done in all of the cancers and showed the PD-L1 expression is linked to a poor prognosis (16, 17). Another meta-analysis has evaluated the prognostic significance of PD-1 and PD-L1 expressions in immune cells of the tumor microenvironment and showed the prognostic role of these molecules in the cancers (18). Four studies in 2018 and 2019, have examined the relationship between PD-L1 expression on tumor cells and prognostic parameters in patients with CRC and concluded that PD-L1 overexpression is related to a poor prognosis (19-22). Most of these studies consist of English-language articles only; and focused on the expression of PD-L1 on tumor cells.

This systematic review and meta-analysis aims to evaluate the association between PD-1/PD-L1 expression on both tumor cells and TIICs and prognostic parameters (e.g., overall survival (OS), progression-free survival (PFS), disease-free survival (DFS), and recurrent-free survival (RFS)) in patients with colorectal cancer. The secondary objective is about evaluating the relationship between PD-1/PD-L1 expression and clinicopathological features (e.g., stage of cancer, tumor location, microsatellite stability status, differentiation grade, vascular or lymphatic invasion).

Based on our knowledge, this study, for the first time, will simultaneously evaluate the association between the expression of both PD-1 and PD-L1 on both tumor cells and TIICs and compare their expression with prognostic parameters in patients with CRC. Moreover, no restriction on publication language will be applied to this study.

## 2. Methods

This protocol has been prospectively registered in the PROSPERO International prospective register of systematic reviews (registration NO. CRD42020156233). The methods adopted for this review conform to Preferred Reporting Items for Systematic Review and Meta-Analysis (PRISMA) checklist for systematic reviews (23), and the protocol has been adopted to the PRISMA-P 2015 checklist (<u>supplemental file 1</u>) (24). Furthermore, the number of included or excluded original studies during the study selection will be described through the PRISMA flow diagram (figure 1) (23).

**Figure 1.**
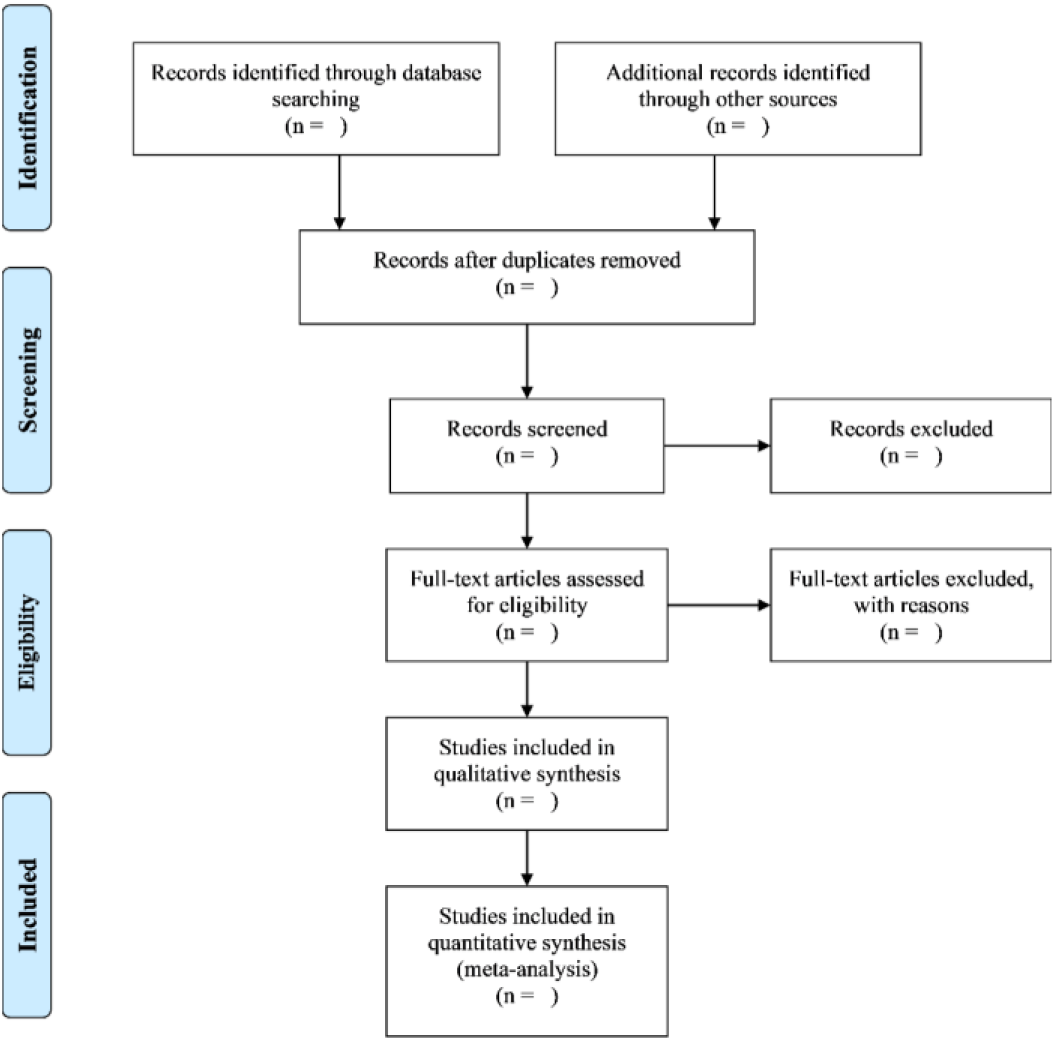
PRISMA flow diagram

### 2.1 Inclusion and exclusion criteria

#### 1. Study characteristics

1. Only cohort studies will be included in this study.
2. There is no restriction on the language of studies (Google Translate will be used to translate non-English articles appropriately).
3. There is no geographic limitation for the studies.
4. Case reports, case series, letters, review articles, and experimental studies will be excluded.

#### 2. Participant characteristics

1. All patients histologically confirmed as having CRC and also had been undergone surgery without preoperative treatment will be included.
2. Patients with sporadic CRC will be included, and patients with hereditary CRC (Lynch syndrome or non-polyposis colon cancer) will be excluded.
3. Studies with any age groups, sexes, and races will be included.
4. All stages of cancer will be included.

#### 3. Exposure

(5) Only studies evaluated PD-1 or PD-L1 expression by immunohistochemistry (IHC) will be included.
(6) Studies with any cut-off values of PD-1 or PD-L1 expression will be included.

#### 4. Outcome

(7) Studies that showed an association between expression of PD-1 or PD-L1 and prognosis (i.e., OS, PFS, DFS, RFS) will be included.
(8) Studies that presented a hazard ratio (HR) with its confidence interval (CI) or adequate data to assess survival parameters will be included.

### 2.2 Search strategy

Electronic databases such as PubMed/MEDLINE, Scopus, Embase, and Web of Science (WoS) will be searched between 1 January 1990 and 31 March 2021 (The search time interval was extended). The search syntax has been combined of MeSH terms, Emtree terms, and free text words of “programmed cell death 1”, “programmed cell death ligand 1”, “colorectal” and “cancer” and their equivalents. The syntax will be developed in PubMed and adopted for other databases.

#### Other resources

Google Scholar will be searched to find additional relevant studies; furthermore, gray literature, including conference papers and the thesis, will be searched. ProQuest database will also be searched for the relevant thesis. Moreover, key journals and also reference lists of included researches will be manually searched for relevant references.

### 2.3 Data collection and analysis

All the records from all databases will be imported to the EndNote X8 software, and duplicated researches will be removed from the library.

#### Study selection

Two reviewers independently will screen all the records attained from the search strategy through titles and abstracts for choosing eligible studies. Afterward, they will independently assess the full text of selected articles at the previous step, according to the eligibility criteria to include relevant studies. The reviewers will discuss to resolve the disagreements, and when it is not attained, a third reviewer will act as a referee.

#### Data extraction

The required data will be independently extracted from original articles by two reviewers. Discrepancies will be resolved by consensus, and when it is not achieved, a third reviewer will act as a referee and decide which data should be entered. The required data for extraction from the included studies, are listed below:

#### 1. Study characteristics

First author’s name, year of publication, the country which study was performed, sample size, and follow-up duration.

#### 2. participants characteristics

Age, gender, stage of cancer, tumor location (right colon, left colon and rectum), microsatellite stability status, differentiation grade (poorly differentiated/ undifferentiated or moderately/ well-differentiated), vascular invasion status, lymphatic invasion status, and type of postoperative treatment (chemotherapy or chemoradiotherapy).

#### 3. Exposure details

Tissue sampling (tissue microarray or whole section), type of antibody used for IHC (monoclonal or polyclonal antibody), cut-off values (for PD-1 and PD-L1 expression), type of cells which express PD-1/PD-L1 (tumor-infiltrating immune cells or tumor cells), the measurement method of mismatch repair deficiency (dMMR) (i.e., IHC, PCR, etc.).

#### 4. Outcome measures

Outcome parameters (OS, PFS, DFS, and RFS), the HRs and their 95% CIs of outcome parameters, the risk ratio (RR) of the relationship between PD-1/PD-L1 expression and clinicopathological characteristics and its 95% CI.

##### Dealing with data in special conditions

In the case of missing necessary data, we will send the email three times in two weeks to the corresponding authors of the eligible studies for explaining the aim of the review and requesting their data. If we do not receive any response, we will try to extract the data from survival curves with <u>WebPlotDigitizer</u> version 4.2 (25); and when it is not possible, we will exclude the study. If one study

has multiple publications, we will use the complete version of it.

##### Risk of bias assessment

Two assessors will assess the risk of bias independently, according to the Newcastle-Ottawa Scale (NOS) for cohort studies, which has three domains: selection (4 items), comparability (1 item), and outcome (3 items). Questions that receive a star will calculate in the final score (26, 27). Conflicts between the assessors will be resolved by consensus, and if it is not possible, a third assessor will act as an arbitrator.

### 2.4 Statistical analysis and data synthesis

All analyses will be performed on the aggregated data extracted from primary studies using Stata software version 13 (StataCorp LP, College Station, Tx, USA). The meta-analysis will be used to assess the prognostic significance of PD-1/PD-L1 expression in patients with CRC. If there is a sufficient number of primary studies, meta-analysis will be conducted, and if it is not applicable, meta-synthesis may be applied. According to the study conditions, either the random effect model (REM) or fixed effect model (FEM) will be used. Time-to-event (survival) data will be analyzed using the HR measure with its 95% CI. Besides, the RR measure with its 95% CI, will be used to analyze dichotomous data. Moreover, forest plots will be utilized to present the data from the meta-analyses. A *P*-value ≤ 0.05 will be considered as statistical significance.

#### Assessment of heterogeneity

Statistical heterogeneity will be assessed by Cochran’s Q (𝒳^2^) test with related P-value, and the I^2^ statistic as recommended by the Cochrane Handbook for Systematic Reviews of Interventions (28). The I^2^ statistic will be interpreted by the following guide (29):

- “0% to 40%: not important;
- 30% to 60%: moderate heterogeneity;
- 50% to 90%: substantial heterogeneity;
- 75% to 100%: considerable heterogeneity.”

In the case of considerable heterogeneity (I^2^ > 75%), either subgroup analysis or meta-regression will be conducted. Likely sources of heterogeneity would be tissue sampling, follow-up period, PD-1/PD-L1 cut-off point, country, and quality of studies.

#### Assessment of publication bias

Publication bias will be assessed by “funnel plot”, “Begg’s statistical test” and “Egger’s statistical test and graph” (30, 31). If the above methods show evidence of publication bias, the “Trim and Fill method”(32, 33) will be used to correct the effect. A *P* ≤ 0.05 will be defined as statistical significance.

#### Sensitivity analysis

Sensitivity analysis will be performed by One-study remove (leave-one-out) method. If one of the combinations shows different results from others, the characteristics of that study will be accurately assessed.

## Results

This is an on-going study, and according to the meta-analysis of the aggregated data from the relevant primary studies, the association between PD-1/PD-L1 expression and prognostic parameters (such as OS and DFS) will be reported.

## Discussion

PD-1 and PD-L1 are two immune checkpoint molecules that regulate immune responses. According to the role of these molecules in suppressing the immune system, several studies have been performed to evaluate the prognostic significance of these markers in various types of cancers (10, 11, 34). However, there is controversy on the prognostic role of PD-1 and PD-L1 in patients with CRC (12-15). In the current systematic review and meta-analysis, we will comprehensively search the databases and pool the aggregated data to evaluate the prognostic impact of PD-1 and PD-L1 in CRC. The results of this study will be published in a peer-reviewed journal.

The strengths of this study are: prospectively protocol registration, searching ProQuest database, searching gray literature and key journals, and no limitation in the language of studies.

## Data Availability

The data that support this study are available on request from the corresponding author. The data are not publicly available due to the ongoing process of this study.

## Author contribution

FA and SS designed the study, prepared the draft of the protocol, and developed the search syntax. LJ and MM revised the manuscript. Screening and selection of the primary studies will be done by FA and SS under the guidance of MM. Data extraction will be done by FA and HB under the supervision of MM. The risk of bias will be assessed by FA and AKh under the superintendence of LJ. LJ also will analyze the data extracted from the included studies. All the authors will revise the final version of the manuscript before the publication.

